# Clinical utility of fluid biomarkers in the evaluation of cognitive impairment: a systematic review and meta-analysis

**DOI:** 10.1101/2022.07.06.22277146

**Authors:** Jemma Hazan, Michelle Wing, Kathy Liu, Suzanne Reeves, Robert Howard

**Author notes:** **Corresponding author details:** Dr Jemma Hazan, Division of Psychiatry, University College London, Maple House, 149 Tottenham Court Road, London W1T 7BN London, UK. **Sponsor:** none.

## Abstract

**Background:** The analytical and clinical validity of fluid biomarkers, measured typically in cerebrospinal (CSF) serum or plasma, has been extensively researched in dementia. Further work is needed to assess the ability of these biomarkers to improve diagnosis, management, and health outcomes in the clinical setting

**Objectives:** To assess the added value and clinical utility of fluid biomarkers in the diagnostic assessment of cognitively impaired patients under evaluation for Alzheimer’s disease (AD).

**Methods:** Systematic literature searches of Medline, EMBASE, PsycINFO, Web of Science research databases were conducted on 17^th^ December 2022. Data from relevant studies were extracted and independently screened for quality using a tool for bias. Clinical utility was measured by clinicians’ changes in diagnosis, diagnostic confidence, and patient management (when available), after their examination of patients’ fluid biomarkers. Cost-effectiveness was assessed by consideration of additional cost per patient and quality-adjusted life years (QALY).

**Results:** Searches identified 18 studies comprising 2199 patient participants and 599 clinicians. The meta-analysis revealed that clinicians’ use of fluid biomarkers resulted in a pooled percentage change in diagnosis of 25% (95% CI: 14–37), an increase in diagnostic confidence of 14% (95% CI: 9–18) and a pooled proportion of patients whose management changed of 31% (95% CI 12–50). CSF biomarkers were deemed cost effective, particularly in memory services, where pre-test AD prevalence is higher compared to a primary care setting.

**Conclusions:** Fluid biomarkers can be a helpful additional diagnostic tool for clinicians assessing patients with cognitive impairment. In particular, CSF biomarkers consistently improved clinicians’ confidence in diagnosing AD and influenced on diagnostic change and patient management. Further research is needed to study the clinical utility of blood-based biomarkers in the clinical setting.

## Introduction

There are over 850,000 people with dementia in the UK^1^, and numbers are expected to rise as the population ages, with over 1.1 million people living with AD in the UK by 2025. The diagnosis of Alzheimer’s disease (AD), which is the most common form of dementia, has advanced in the last decade through the availability of in vivo biological measures or “biomarkers”. These biomarkers can detect the pathological hallmarks of AD: pathological tau and beta amyloid proteins, as well as neurodegeneration^2^. Biomarkers have been incorporated into the diagnostic framework of AD in clinical research^3^. A consensus “roadmap” was set out in 2017 to aid the incorporation of these biomarkers into the clinical setting to improve diagnostic accuracy^4^.

Fluid biomarkers comprise cerebrospinal fluid (CSF) and blood plasma or serum measures. CSF biomarkers are currently used in the diagnostic investigation of AD at specialist tertiary Neurology Centres in the UK and have demonstrated analytical validity^5,6^. Validated AD CSF biomarkers include amyloid-β1-42, total-tau, and phosphorylated-tau181 (ptau-181)^7^. A reduction in Aβ42 and raised levels of ptau are indicative of Aβ and tau pathologies in AD, while increased total-tau is a non-specific marker of neuronal injury^8^. Many studies have demonstrated the correlation between CSF levels and neuropathology^9 10^. There have been recent advancements in the validation of blood-based biomarkers, such as ptau-181 and ptau-217, which have been shown to have similar sensitivity and specificity to CSF-biomarkers^11 12 13^. The level of neurofilament light (NfL) is raised in neurological diseases which cause axonal damage such as AD and frontotemporal dementia (FTD)^14 15^. These fluid biomarkers have garnered attention as, in contrast to imaging biomarkers such as amyloid PET, they are cheap, quick, and simple to obtain in a clinical setting^16^.

Biomarkers may also assist clinicians in differentiating AD from non-AD dementias, and mild cognitive impairment (MCI) from early AD^17 18 19^.

There have been several studies exploring the validity and diagnostic accuracy of fluid biomarkers in AD^20^. However, to date there has been no systematic review of the clinical utility of fluid biomarkers in the diagnostic evaluation of cognitively impaired patients. In this study we aim to assess the real world added value and clinical utility, defined as relative improvement in clinicians’ diagnostic confidence or change in diagnosis or management, of fluid biomarkers in patients being evaluated for cognitive impairment due to AD.

## Methods

### Study design

A systematic review with mixed methods quantitative and narrative synthesis was conducted following the Preferred Items for Reporting of Systematic Reviews and Meta-Analyses (PRISMA) guidelines^21^.

### Eligibility Criteria

Included studies performed a diagnostic and clinical utility analysis of CSF or blood biomarkers, where clinicians cognitively assessed at least 10 cognitively impaired participants of any age undergoing evaluation for AD. Peer-reviewed published studies in English, were included if their primary or secondary outcome included at least one of change in diagnosis, diagnostic confidence, patient management or cost analysis. We excluded reviews, protocols, and conference presentations.

### Search Strategy

An online literature search was carried out on the 17^th^ December 2021 using Medline, Embase, PsycINFO and Web of Science (WoS) databases, using the terms listed in the Supplementary Data Appendix 1. The search terms were modified to meet the criteria for medical subject headings in the various databases. The references of identified articles were also screened to ensure all relevant studies were included.

### Data Extraction

Two authors (JH and MW) independently screened and selected potentially relevant abstracts and assessed the full study texts according to eligibility criteria. Any disagreement between authors was resolved through discussion. If there was any further disagreement this was resolved by discussion with a third author (SR).

Two authors (JH & MW) independently extracted data. If there was a disagreement this was resolved in discussion between the two raters. We extracted data on study characteristics (design, setting, duration, intervention, inclusion criteria), relevant outcomes (change in diagnosis, diagnostic confidence and change in management plan), and participant characteristics (population, sample size, initial diagnosis, age, sex, demographics).

### Risk of Bias in individual studies and Quality Assessment

Two authors (JH and MW) independently assessed studies for bias using a modified Quality Assessment Tool for Quantitative Studies, originally developed by the Effective Public Health Practice Project (EPHPP)^22^. Any disagreement was resolved by discussion with a third author (SW). Quality of studies was assessed across several domains including selection bias, study design, confounders, data collection method and withdrawals and dropouts. The scores were collated to give an overall global rating for each paper as “Strong”, “Moderate” or “Weak”. If a study received a weak rating in all areas of bias, it was excluded from the review.

### Synthesis of results and meta-analysis

A mixed methods quantitative and narrative synthesis was carried out due to the small number of studies and heterogeneity in study methodology.

In terms of the quantitative analyses, the percentage of change in diagnoses, diagnostic confidence and management was computed using available study data. A random-effects meta-analysis was conducted to calculate pooled estimates of the percentage change in diagnoses, confidence, and management, due to the heterogeneity in study settings and study populations^23^. Sub-analyses were performed on the percentage change in AD diagnoses, i.e., changes in diagnosis from AD to non-AD and from non-AD to AD. The I^2^ statistic was used to assess the degree of heterogeneity of the percentage change in diagnosis, confidence, and management across studies^24^. We followed Tu, YK 2016 methodology for testing the relationship between percentage change and baseline values, which uses a modified Pearson’s test^25^. All analyses were performed using R Software R version 4.1.2; R Foundation for Statistical Computing^26^.

## Results

### Study Identification

18 studies were identified for inclusion. The PRISMA flowchart is displayed in Figure 1

**Fig. 1.**
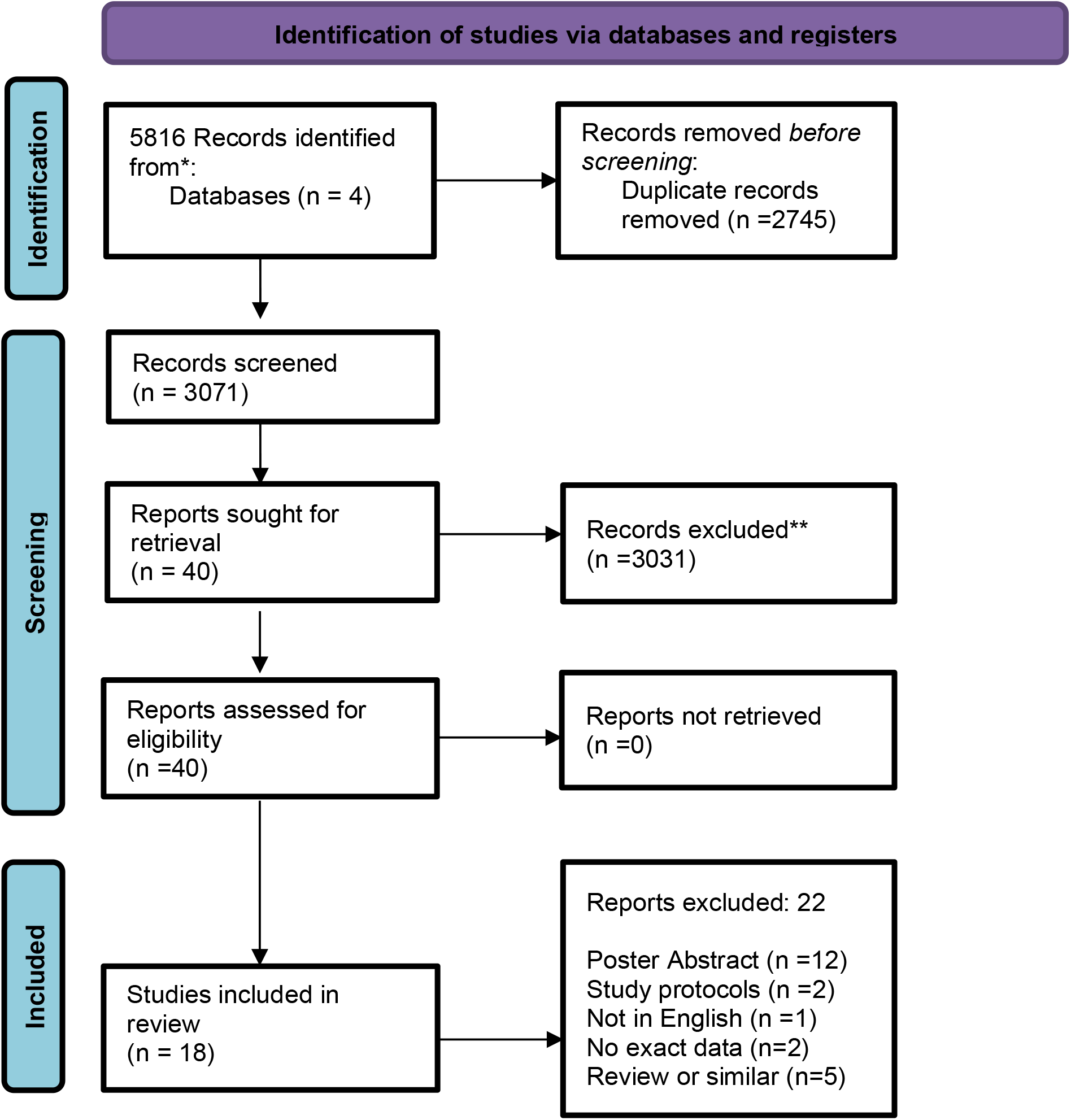
Preferred Reporting Items for Systematic Reviews and Meta-Analyses (PRISMA) flowchart of selected papers

### Study characteristics

Study characteristics are shown in Table 1, Table 2, and Table 3.

**Table 1:** Table of Study Characteristics

| Table 1: Study characteristics |  |  |  |  |  |  |  |  |
| --- | --- | --- | --- | --- | --- | --- | --- | --- |
| Author, Year of Publication | Country | Type of study | Referral setting | Intervention | Study Duration | Patient sample size (N) | Clinician sample size (N) | Patient inclusion criteria |
| Balasa et al, 2013 | Spain | Prospective observational Study | Specialist Outpatient Clinic | LP for CSF Biomarkers: Aβ42, t-tau, and p-tau | 2009-2013 | 157 | NR | patients < 65 years |
| Boelaarts et al, 2020 | The Netherlands | Prospective observational Study | Community geriatric outpatient clinic | LP for CSF Biomarkers: Aβ42, t-tau, and p-tau | 2010-2016 | 69 | NR | Patients <71 years |
| Cognat et al, 2019 | France | Prospective observational Study | 29 Secondary & Tertiary memory clinics | LP for CSF Biomarkers: Aβ42, t-tau, and p-tau | 2012-2014 | 153 | 128 | Initial diagnosis of MCI |
| Duits et al, 2015 | The Netherlands | Prospective observational Study | Memory clinic | LP for CSF Biomarkers: Aβ42, t-tau, and p-tau | 2011-2012 | 351 | 5 | All patients visiting the clinic |
| E.M. van den Brink et al, 2020 | Canada | Retrospective observational study | Specialist Memory Clinic | LP for CSF Biomarkers: Aβ42, t-tau, and p-tau & FDG-PET | 2017-2019 | 28 | NR | Atypical dementia presentations, |
| Falgas et al, 2019 | Spain | Prospective observational study | Specialist Memory Clinic | Patients underwent biomarkers which included: MRI, LP for CSF Biomarkers: Aβ42, t-tau, and p-tau, FDG-PET, Amyloid PET | not reported | 40 | 5 | <65 years of age , MMSE score ≥ 18 |
| Gjerum et al, 2021 | Denmark | Retrospective observation incremental study | Memory Clinic | Patients randomised to addition of either 2-[18F]FDG-PET or CSF Biomarkers: Aβ42, t-tau, and p-tau | 2015-2016 | 81 | 2 | Cognitive impairment due to neurodegenerative disease, (MMSE ≥ 18, CDR ≥ 1.0, undergone T1-weighted MRI ≥ 1.5 Tesla, addition of CSF biomarkers deemed useful by clinician |
| Gooblar et al, 2015 | USA | study using simulated clinical vignettes | Primary, Secondary and Tertiary Centres | Simulated CSF values | January-July 2013 | 2 | 193 | 2 Simulated clinical vignettes |
| Handels et al, 2017 | The Netherlands | Cost-effectiveness analysis | NA | Simulated CSF values | NA | NA | NA | Simulated patients with MCI |
| Kester et al, 2010 | The Netherlands | Prospective observational Study | One local hospital memory clinic | LP for CSF Biomarkers: Aβ42, t-tau, and p-tau | 2005-2008 | 109 | 2 | All patients visiting the clinic |
| Lee et al, 2017 | Canada | Cost-effectiveness analysis | NA | Simulated CSF values | NA | NA | NA | Simulated patients with suspected AD |
| Mouton-Liger et al, 2014 | France | Prospective observational study | secondary or tertiary memory centres | LP for CSF Biomarkers: Aβ42, t-tau, and p-tau | not specified | 561 | 128 | When a clinician considered a patient eligible for CSF biomarkers |
| Paquet et al, 2016 | France | Prospective observational Study | 29 Secondary & Tertiary memory clinics | LP for CSF Biomarkers: Aβ42, t-tau, and p-tau | 2014-2016 | 69 | 128 | Initial main diagnosis of a psychiatric disorder |
| Ramusino et al, 2019 | Italy | Prospective Study | multicentre, 4 Memory Clinics | LP for CSF Biomarkers: Aβ42, t-tau, and p-tau & Amyloid-PET | 2015-2017 | 71 | 2 | MCI or mild dementia possibly due to AD, Age range 55-90 years, score < 4 on the modified Hachinski ischemic scale At least 5 years of education |
| Stiffel et al, 2021 | Canada | Prospective observational Study | A tertiary care memory clinic | LP for CSF Biomarkers: Aβ42, t-tau, and p-tau | 2015-2020 | 262 | NR | participants included if basic tertiary care work up, neuropsychological evaluation and FDG-PET did not provide conclusive diagnosis |
| Valcarcel-Nazco et al, 2014 | Spain | Cost-effectiveness analysis-Economic evaluation | NA | Simulated CSF values | NA | NA | NA | Simulated patients with MCI & other dementias |
| Willemse et al, 2021 | The Netherlands | Prospective observational Study | A tertiary care memory clinic | Serum Neurofilament Light (sNfL) | May-October 2019 | 109 | 6 | All patients visiting the clinic |
| Ye et al, 2021 | China | Prospective observational Study | Neurology Clinic | LP for CSF Biomarkers: Aβ42, t-tau, and p-tau | 2015-2019 | 137 | not reported | All patients visiting the clinic |
| Abbreviations: MMSE - Mini Mental State Examination; CDR- Clinical Dementia Rating; LP-Lumbar Puncture; CSF- cerebrospinal Fluid; PET FDG- Fluorodeoxyglucose positron emission tomography; Aβ42- Amyloid Beta 42; T-tau- Total Tau; P-tau- Phospho-tau; AD- Alzheimer's Disease; NR- Not Reported | | | | | | | | |

**Table 2:**
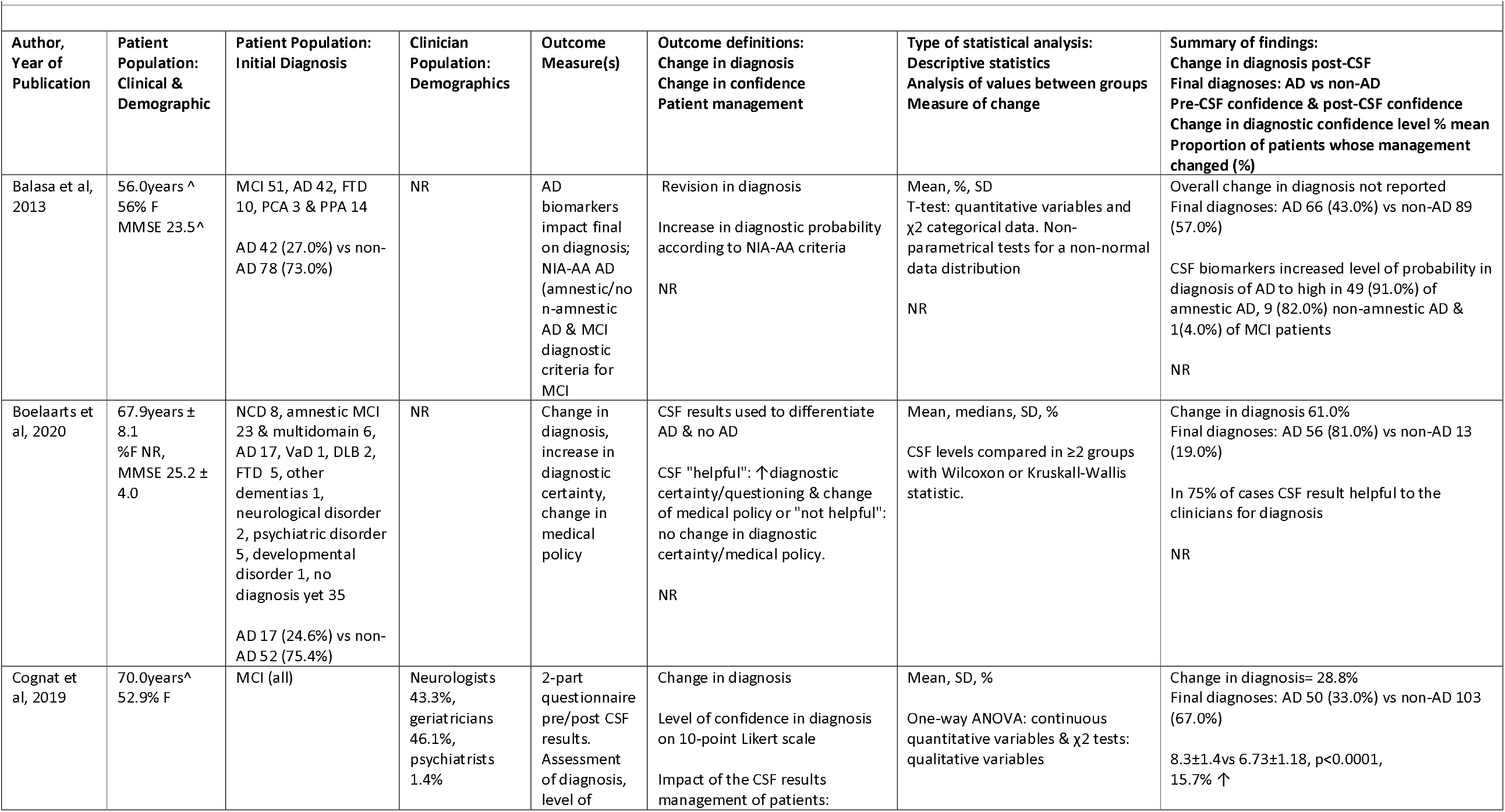
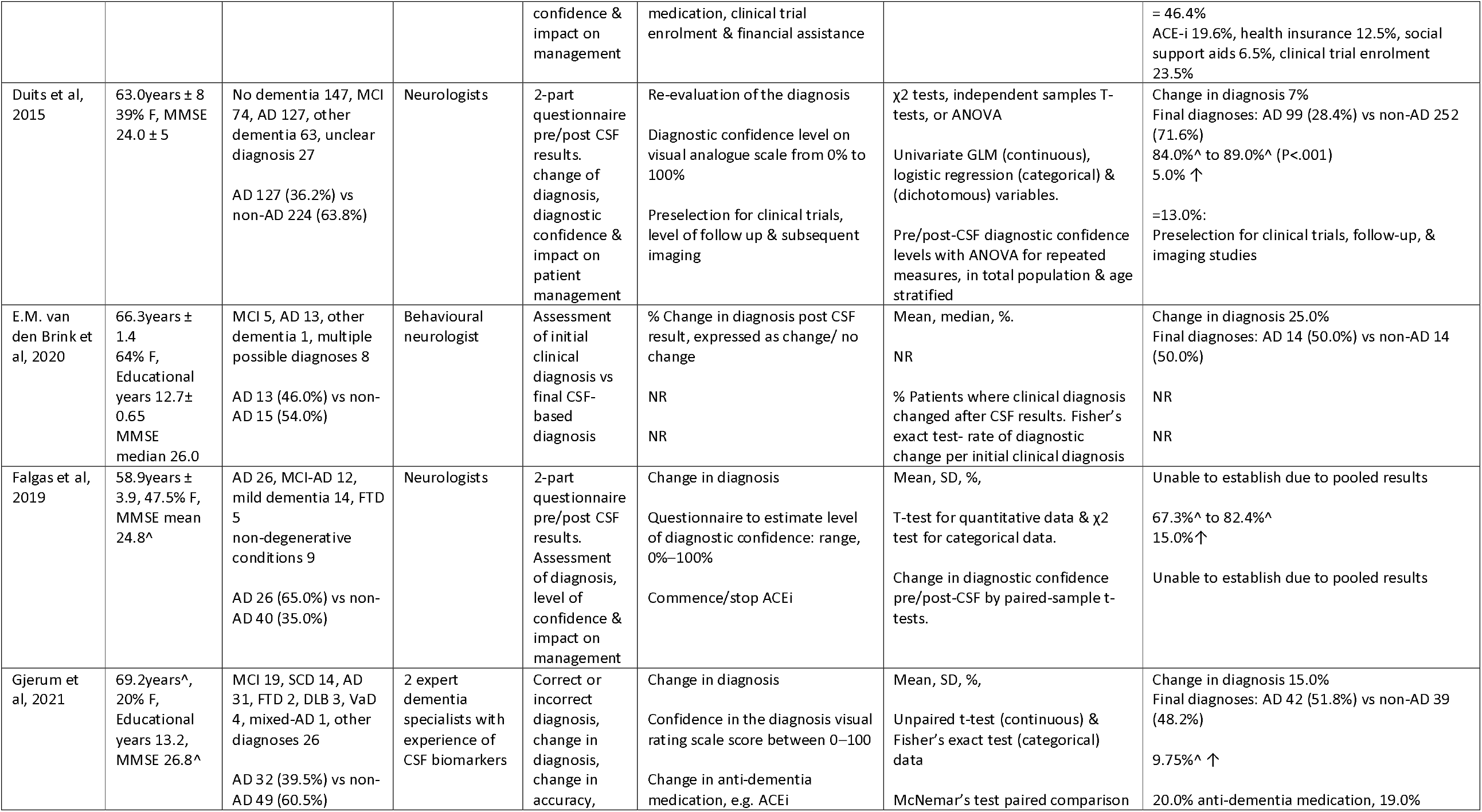
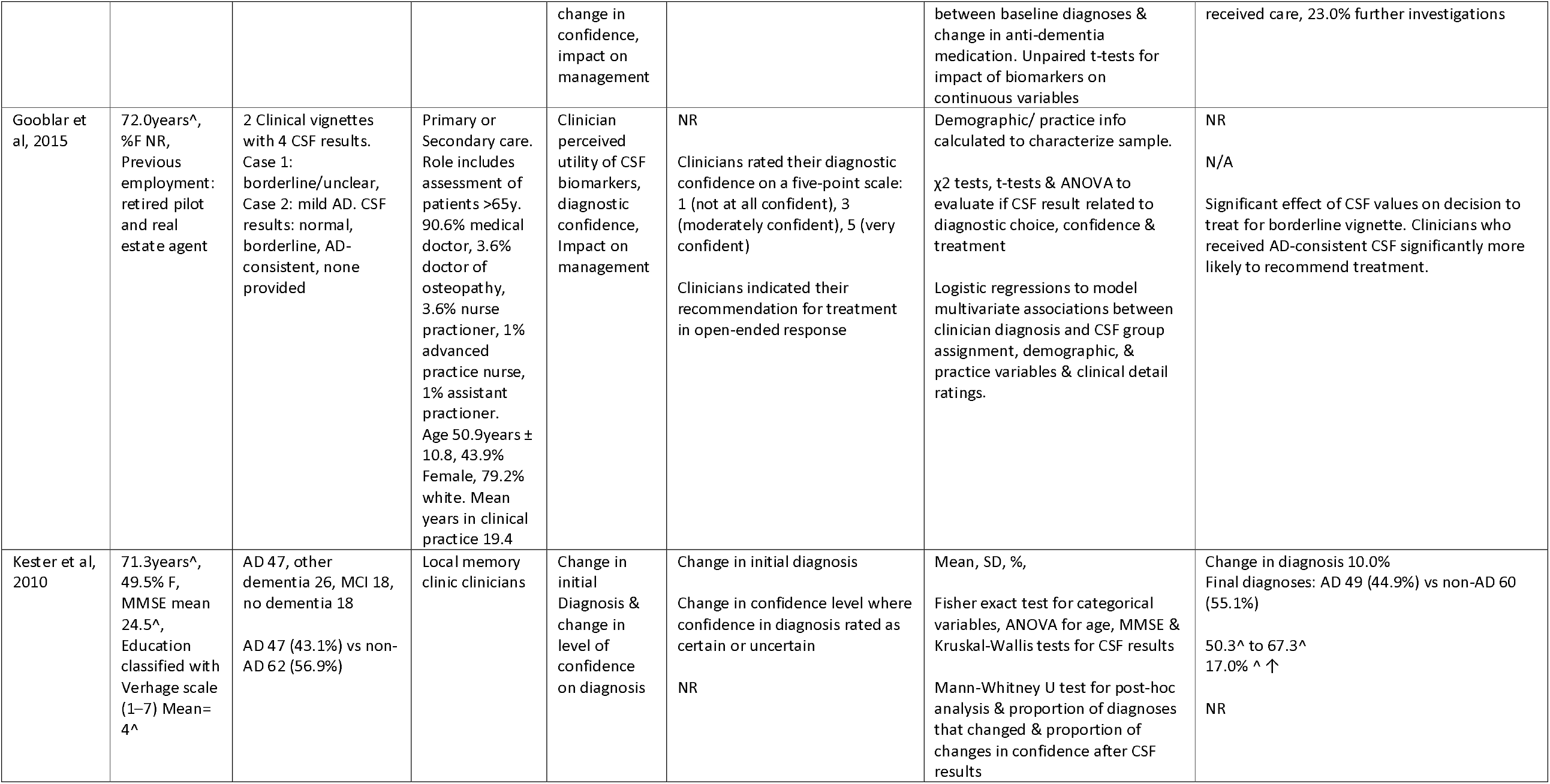
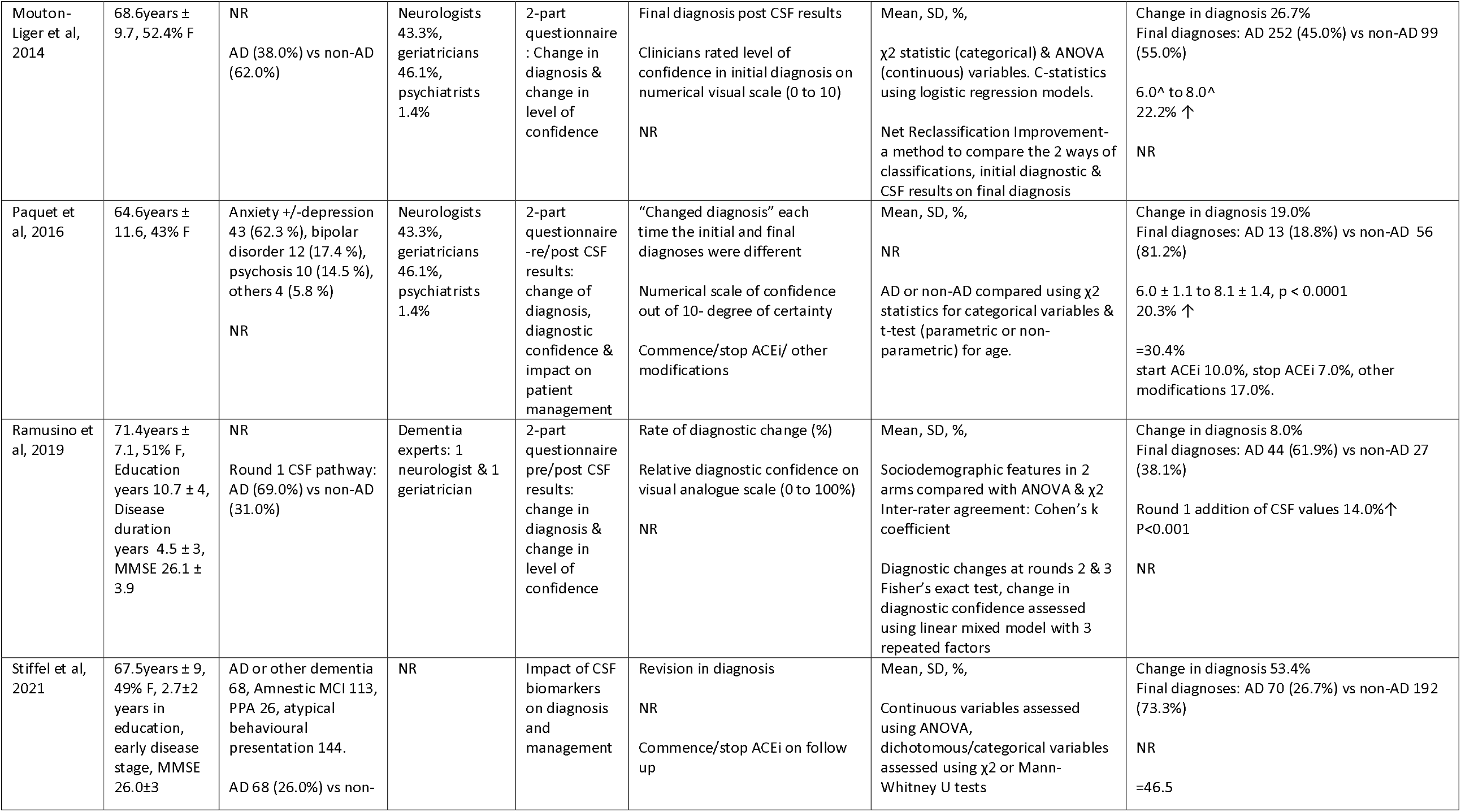
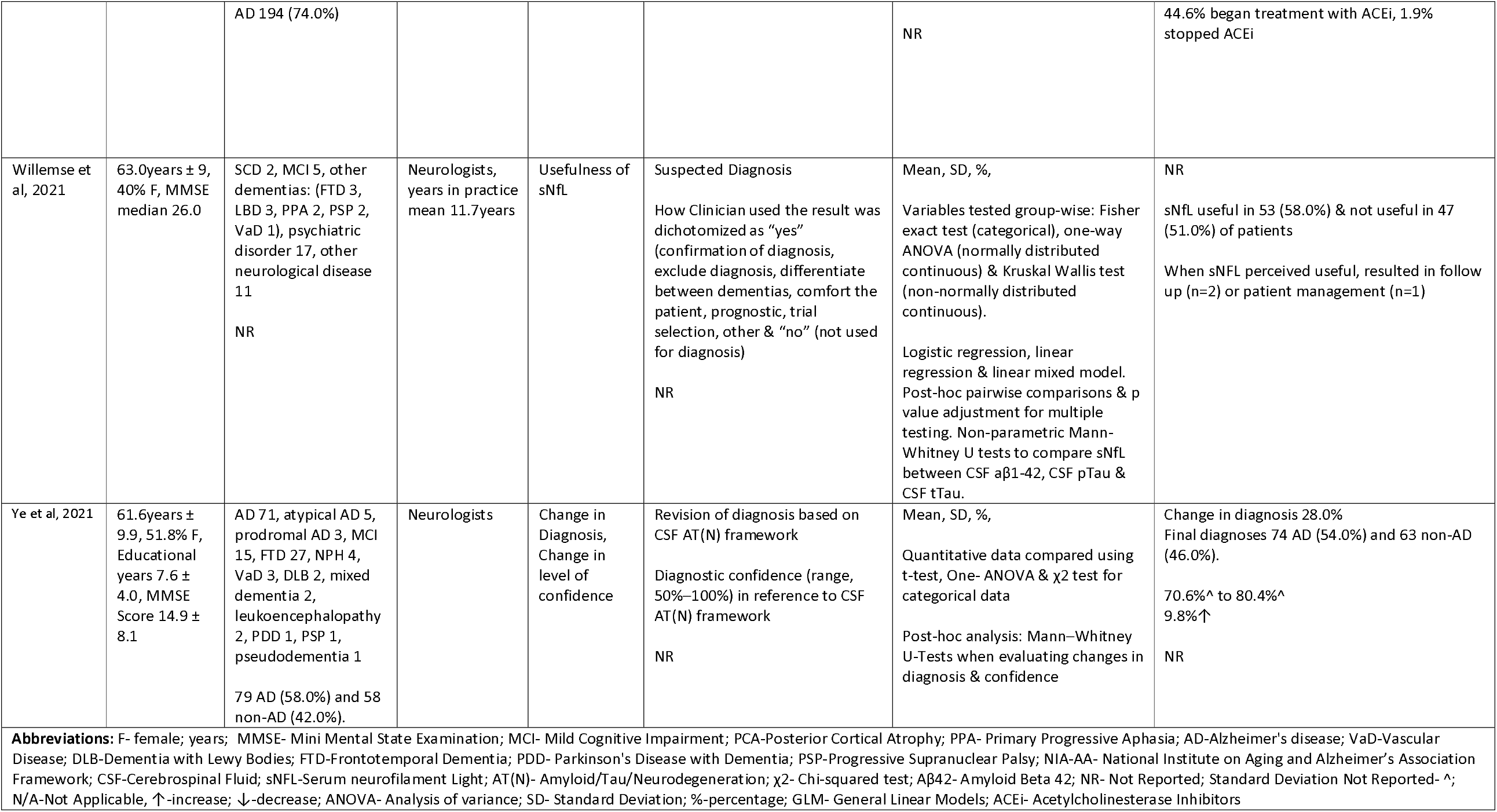
Table of Study Characteristics & Findings- Observational Studies

**Table 3:** Table of Study Characteristics & Findings- Cost-effectiveness Analysis Studies

| Table of Study Characteristics & Findings: Cost-effectiveness Analysis |  |  |  |  |  |
| --- | --- | --- | --- | --- | --- |
| Author, Year of Publication | Type of study | Aim | Outcome measure | Statistical analysis | Summary of findings |
| Handels et al, 2017<br>The Netherlands | Cost-effectiveness analysis | To estimate the potential ICER of adding CSF biomarker testing to the standard diagnostic workup to determine the prognosis for patients with MCI | Accuracy of prognosis<br>QALY<br>Additional cost per patient<br>ICER | Simulated data model using a merged dataset | Improved the accuracy of prognosis by 11%<br>Average QALY gain of 0.046<br>€432 additional costs per patient<br>ICER of €9416 per QALY gained |
| Lee et al, 2017<br>Canada | Cost-effectiveness analysis | To estimate the lifetime costs & QALYs of CSF biomarker analysis in a cohort of patients referred to a neurologist or memory clinic with suspected AD who remained without a definitive diagnosis of AD or another condition after neuroimaging | Additional cost per patient<br>QALY<br>ICER | Markov Model | AD pre-test probability of 12.7%:<br>Average QALY gain of 0.015<br>ICER of \$11,032 per QALY gained<br>\$165 additional costs per patient, |
| Valcarcel-Nazco et al, 2014<br>Spain | Cost-effectiveness analysis- Economic evaluation | To determine the cost-effectiveness of the CSF biomarkers to diagnose AD in MCI & dementia patients. | Cost & effectiveness | Probabilistic sensitivity analysis using 2nd order Monte Carlo simulations. Acceptability curves were calculated and ANCOVA models applied to simulation results | MCI Patients: lower average cost per patient of €1832.65<br>AD patients: higher average cost per patient of €1133.82<br>Dominant ICER for MCI patients |
| Abbreviations: ANCOVA- Analysis of Covariance; AD- Alzheimer's Disease; MCI- Mild Cognitive Impairment; ICER-Incremental Cost Effectiveness Ratio; QALY-Quality-adjusted life year; CSF- Cerebrospinal Fluid |  |  |  |  |  |

### Study design

Most (13 of 18, 72%) of the included studies were prospective observational studies ^27–38^., One study was a survey of clinicians using simulated clinical scenarios ^44^. Other study designs included three cost-effectiveness analyses ^39–41^ and two retrospective observational studies ^42,43^. Most studies (14 of 18, 78%) assessed the impact of CSF biomarker results, and only one study included blood-based biomarkers^37^. The mean sample sizes of patient and clinician participants were 147 and 60 respectively. Eight studies were performed at specialist memory clinics with a single site, five were multi-centre and two were early-onset dementia clinics.

### Patient and clinician characteristics

The mean age of patient participants in the studies was 66.1 years (+/- SD 4.73), and two studies restricted inclusion criteria to patients aged < 65 years^27,31^..

Of the 18 studies, AD and MCI were the most common initial (pre-biomarker) diagnoses. The initial (pre-biomarker) diagnoses for patient participants in the studies included: Subjective cognitive disorder (SCD), MCI, AD dementia, frontotemporal lobe dementia (FTLD), vascular dementia (VaD), dementia with Lewy bodies (DLB), dementia with unknown aetiology, Parkinson’s disease and Parkinson’s Plus Syndromes, psychiatric disorders or “other”. In addition to AD, the final (post-biomarker) dementia diagnoses included progressive supranuclear palsy, Creutzfeldt-Jakob disease (CJD), Corticobasal degeneration and Huntington’s disease. Non-demented patients were categorized as “no cognitive disorder”, symptoms of a cognitive disorder caused by a developmental disorder, or a psychiatric or neurological disorder.

Most studies recruited clinicians with a specialty in Neurology, but only one study provided detailed clinician demographics^44^

### Outcome measures

Outcome measures are described in Table 2. The majority (14 of 18, 78%) of studies required the clinician to complete a pre- and post-CSF results questionnaire, listing initial and final diagnoses respectively^27–31,33–38,43,44^. Of these studies, eight examined how CSF biomarker results changed diagnostic confidence ^29–33,38,43,44^, and seven assessed their impact on patient management, defined as the initiation or discontinuation of dementia medications such as cholinesterase inhibitors, the ability to enrol in clinical trials, and/or length of follow-up ^29–31,34,36,43,44^.

Of the three studies that performed a cost-effectiveness analysis of CSF biomarkers to diagnose AD ^39–41^, two examined lifetime costs and quality-adjusted life-years (QALYs) ^39,40^ and one performed an economic evaluation^41^. Outcome measures are described in Table 3.

### Quality assessment

Most (15 of 18, 83%) studies were assessed as being of moderate quality^27–38,42–44^ and three studies were assessed as being of high quality^39–41^. No studies were assessed to be of low quality (Supplementary Table 1).

### Findings

#### Change in Diagnosis

Eleven studies, comprising 1891 patient participants and 395 clinician participants, reported on the percentage change in clinicians’ diagnosis after the availability of fluid biomarker results ^28–30,32–36,38,42,43^, which ranged between 7–61%. The overall pooled percentage change in diagnoses was 25% (95% CI: 14–37) and there was substantial heterogeneity (I^2^ 97%, p < 0.001) (Fig. 2a). Subgroup analyses found no significant change in diagnoses from initial AD to final non-AD or initial non-AD to final AD (Supplementary material Figure 1a & 1b).

**Fig. 2.**
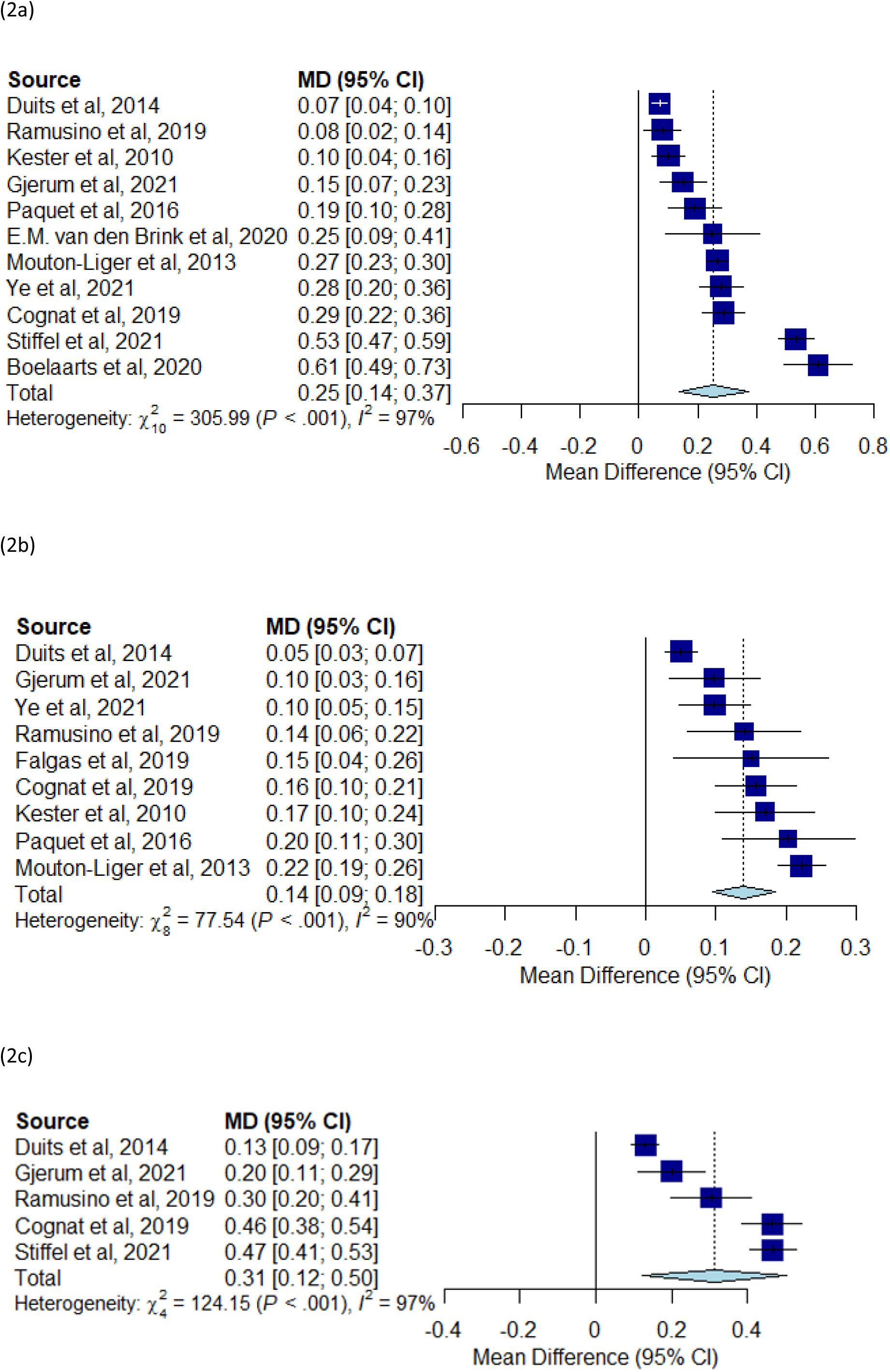
(a) Forest plot showing the pooled percentage change in diagnosis. (b) Forest plot showing the pooled percentage change in confidence. (c) Forest plot showing the pooled proportion of patients whose management changed (%).

Two studies explored the accuracy of clinicians’ final diagnoses through longitudinal patient follow up. In one study, after 12-months of follow up, 89% of patients’ diagnoses were found to be correctly classified ^43^. Similarly, another study showed that after a mean follow up time of 31 months, 88% of AD participants maintained their diagnosis and all MCI participants who had positive CSF results progressed to AD dementia^27^.

#### Clinician Rated Diagnostic Confidence

Eight studies calculated and showed an overall increase in diagnostic confidence ^29–35,38^, ranging from 5 to 22% (Table 2). The overall pooled percentage change in confidence was 14% (95% CI: 9–18) and there was substantial heterogeneity (I^2^ 88%, p < 0.001) (Fig. 2b). The change in confidence was inversely proportional to the initial pre-test confidence level, such that lower pre-test confidence was associated with a greater percentage change in confidence (Pearson’s r =-0.91, p<0.001).

Two studies that reported an overall increase pooled percentage change in confidence also showed a decrease in diagnostic confidence for a minority of clinicians^30 32^. This was often associated with patients for whom CSF results did not alter the final diagnosis and who had a pre-biomarker diagnosis of subjective memory complaint or a psychiatric disorder ^30^.

#### Change in Management

Five studies, comprising 918 patients, evaluated the impact on CSF biomarkers on patient management ^29,30,35,36,43^. The overall proportion of patients whose management changed after availability of fluid biomarkers ranged between 13-47%, and the overall pooled proportion of patients whose management changed was 31% (95% CI 12–50) with substantial heterogeneity (I^2^: 97%, p < 0.001) (Fig. 2c). The most common management change was the commencement or stopping of cholinesterase inhibitors or other dementia medications (4 of 5 studies).

#### Cost-Effectiveness

Three studies analysed the cost-effectiveness of CSF biomarkers to diagnose AD in MCI and dementia populations: a 2014 Spanish study and two studies published in 2017 from Canada and The Netherlands. All three studies assessed CSF biomarkers (amyloid-beta 1-42, total tau, and phosphorylated tau). In one study^41^, CSF biomarkers were reported to be an alternative, less expensive and more efficient diagnostic tool compared to standard diagnostic procedures in MCI patients, as per guidelines of the National Institute of Neurological and Communicative Disorders and Stroke and the Alzheimer’s Disease and Related Disorders Association(NINDS-ADRDA guidelines). The same study reported that for dementia patients, despite higher uncertainty, CSF biomarkers were also a cost-effective alternative compared to standard clinical diagnostic criteria.

A second study^40^ utilised a Markov model to estimate the lifetime costs and QALYs of CSF biomarkers in patients referred for cognitive assessment with suspected AD, where diagnosis remained unclear after neuroimaging. The study reported that the cost-effectiveness of CSF biomarkers depended on the prevalence of AD in the population, such that with a pre-test probability of AD of 12.7%, the addition of CSF biomarkers to neuroimaging had an incremental cost-effectiveness ratio (ICER) of $11,032 per QALY gained. However, at a lower prevalence such as in the general practice setting, CSF biomarkers were unlikely to be cost-effective at a willingness-to-pay threshold of $50,000 per QALY gained. The study concluded that CSF biomarkers are likely to be cost effective in specialist memory clinics where pre-test prevalence may be greater than 15%.

The third study^39^ found that the use of CSF biomarkers in an MCI population resulted in an ICER of €9,416, although there was a high degree of uncertainty. This was due to the uncertainty of input parameters computed in the model such as expert opinions and risk prediction coefficients.

## Discussion

Previous systematic reviews have reported on the analytic or clinical validity of fluid biomarkers, or the clinical utility of imaging biomarkers such as amyloid PET^45–48^. In this review, we focussed specifically on the clinical utility of fluid biomarkers in the assessment of patients with cognitive impairment and the cost-effectiveness of CSF biomarkers for AD. Use of CSF biomarkers resulted in a change in diagnosis in 25% of cases, although this was not specific to any direction of diagnostic change (AD to non-AD or non-AD to AD). This result is similar to the overall change of diagnosis of 35.2% after amyloid-PET ^45^.

Biomarker results are likely to provide an additional diagnostic assessment tool that clinicians will consider in combination with clinical findings. In one study that provided simulated clinical vignettes to clinicians^44^, an AD clinical presentation with AD CSF results led to a significantly increased odds of an AD diagnosis, whereas when clinicians were given borderline CSF values, they relied on other clinical information to decide on the final diagnosis. Also, when clinicians were shown a mild AD clinical presentation with normal CSF results, they often chose a diagnosis of unknown aetiology, and when clinicians were shown an ambiguous clinical presentation with AD CSF result, they were more likely to make an AD diagnosis.

Studies consistently reported that CSF biomarkers improved clinicians’ diagnostic confidence with a pooled mean increase of 14%. This is in comparison to the impact of amyloid-PET where the change in confidence level reportedly ranged from 16 to 44%^45^. However, some studies reported that biomarker results resulted in a reduction in confidence, for example, in the context of unexpected biomarker results when patients with subjective cognitive complaint or a psychiatric disorder had abnormal dementia biomarkers, or when patients initially diagnosed with AD had normal biomarkers. It is relevant that higher diagnostic confidence may not always equate to greater clinical utility^28^, as decreased confidence after CSF results could sometimes help a clinician to question their pre-CSF diagnosis and prevent an incorrect diagnosis. A reduction in diagnostic confidence may also spur further diagnostic tests and have a substantial impact on management.

Use of fluid biomarkers led to a change in management in 31% of cases, mostly involving the initiation or discontinuation of cholinesterase inhibitors. One review examining the impact of amyloid-PET found that the overall change in management was 59.6%^45^, which represents a larger pooled effect size compared to CSF biomarkers. This may be due to factors such as the proportion of patients already prescribed medication and degree of diagnostic certainty prior to amyloid-PET imaging. However amyloid-PET is costly, less accessible and provides information solely on amyloid deposition^49^. Cost-effectiveness analyses revealed fluid biomarkers to be a cost-effective alternative to standard diagnostic work-up^39–41^.

### Limitations

The interpretation of the findings is limited by the small number of included studies, small sample sizes, and high methodological heterogeneity. Most included studies were of moderate quality. Study quality limitations included lack of information about baseline clinician demographics and were observational studies.

Only one study reported on clinician demographics such as ethnicity, age, and level of seniority, and most clinicians were neurologists, so it is unknown how these clinician-factors may have influenced the outcome measures, such as degree of diagnostic confidence and familiarity with the use of fluid biomarkers. The extent to which these findings are generalizable to other clinician specialities involved in making dementia diagnoses is also unclear.

The mean age of patients included in these studies was 66.1 years. Future studies should investigate older patients, who are more representative of local memory service populations. Some studies requested clinicians in Memory Services to complete questionnaires on a voluntary basis, which may have introduced a selection bias as clinicians with a higher inclination to use biomarkers and find them useful in clinical practice may have been more likely to respond^29^.

No studies confirmed the final diagnosis with post-mortem brain study findings, so we were unable to assess the overall diagnostic accuracy of fluid biomarkers. Future larger longitudinal studies would be helpful to assess the diagnosis accuracy of these methods.

Only one study assessed the clinical utility of a serum biomarker, sNfL, in the diagnosis of neurodegenerative diseases^37^. Further studies are needed to assess the clinical impact of other blood-based biomarkers.

### Conclusion

Fluid biomarkers can provide additional value in the diagnostic assessment of cognitively impaired patients presenting to memory clinic through changes in clinical diagnoses, improved diagnostic confidence, and changes to patient management. Fluid biomarkers, especially blood-based biomarkers, offer a simple-to-obtain, cost-effective and scalable test to support clinicians in the diagnosis of Alzheimer’s dementia.

## Data Availability

All data produced in the present work are contained in the manuscript

