## Supplementary data for "Clinical utility of fluid biomarkers in the evaluation of cognitive impairment: a systematic review and meta-analysis"

### Medline Search Strategy

1. Dementia/
2. Alzheimer*.mp.
3. Alzheimer disease/
4. (Cogniti* adj2 (impair* or decline* or loss* or disorder* or deteriorat* or dysfunction*)).mp.
5. 1 or 2 or 3 or 4
6. Cerebrospinal fluid/
7. Amyloid beta-Peptides/
8. tau Proteins/
9. (cerebro-spinal fluid* or cerebrospinal fluid* or csf or spinal fluid*).mp.
10. ((blood or plasma) adj3 (biomarker* or marker* or biological marker*)).mp.
11. (biomarker* or marker* or biological marker*).mp.
12. biomarker/
13. (abeta* or ab42 or ab40 or amyloid beta or beta amyloid).mp.
14. (phospho tau* or total tau* or ptau181 or phosphorylated tau or ptau*).mp.
15. neurofilament.mp.
16. or/6-15
17. Diagnos*.mp.
18. Diagnosis/
19. Diagnosis.fs.
20. or/17-19
21. clinical decision rules/
22. cost benefit.tw.
23. Cost-Benefit Analysis/
24. ((clinical or perceived or clinician* or pragmatic or diagnos*) adj5 (impact or utility or useful* or confidence or decision* or benefit*)).mp.
25. or/21-24
26. 5 and 16 and 20 and 25
27. exp animals/ not humans.sh.
28. 26 not 27

### PsychInfo Search strategy

1. Dementia/
2. Alzheimer*.mp.
3. Alzheimer's Disease/
4. (Cogniti* adj2 (impair* or decline* or loss* or disorder* or deteriorat* or dysfunction*)).mp.
5. 1 or 2 or 3 or 4
6. Cerebrospinal fluid/
7. Beta Amyloid/
8. Tau Proteins/
9. (cerebro-spinal fluid* or cerebrospinal fluid* or csf or spinal fluid*).mp.
10. ((blood or plasma) adj3 (biomarker* or marker* or biological marker*)).mp.
11. (biomarker* or marker* or biological marker*).mp.
12. Biological Markers/
13. (abeta* or ab42 or ab40 or amyloid beta or beta amyloid).mp.
14. (phospho tau* or total tau* or ptau181 or phosphorylated tau or ptau*).mp.
15. neurofilament.mp.
16. or/6-15

17. Diagnos*.mp.
18. Diagnosis/
19. or/17-18
20. "clinical judgment (not diagnosis)"/
21. cost benefit.tw.
22. "costs and cost analysis"/
23. ((clinical or perceived or clinician* or pragmatic or diagnos*) adj5 (impact or utility or useful* or confidence or decision* or benefit*)).mp.
24. or/20-23
25. 5 and 16 and 19 and 24

**Embase Search Strategy**

1. Dementia/
2. Alzheimer*.mp.
3. Alzheimer disease/
4. (Cogniti* adj2 (impair* or decline* or loss* or disorder* or deteriorat* or dysfunction*)).mp.
5. 1 or 2 or 3 or 4
6. cerebrospinal fluid/
7. amyloid beta protein/
8. tau protein/
9. (cerebro-spinal fluid* or cerebrospinal fluid* or csf or spinal fluid*).mp.
10. ((blood or plasma) adj3 (biomarker* or marker* or biological marker*)).mp.
11. (biomarker* or marker* or biological marker*).mp.
12. biomarker/
13. (abeta* or ab42 or ab40 or amyloid beta or beta amyloid).mp.
14. (phospho tau* or total tau* or ptau181 or phosphorylated tau or ptau*).mp.
15. neurofilament.mp.
16. or/6-15
17. Diagnos*.mp.
18. diagnosis/
19. Diagnosis.fs.
20. or/17-19
21. clinical decision rule/
22. cost benefit.tw.
23. "cost benefit analysis"/
24. ((clinical or perceived or clinician* or pragmatic or diagnos*) adj5 (impact or utility or useful* or confidence or decision* or benefit*)).mp.
25. or/21-24
26. 5 and 16 and 20 and 25
27. (exp animal/ or animal.hw. or nonhuman/) not (exp human/ or human cell/ or (human or humans).ti.)
28. 26 not 27

**Web Of Science Search Strategy**

1. TS=(Dementia OR Alzheimer*)
2. TS=((Cogniti* near/2 (impair* OR decline* OR loss* OR disorder* OR deteriorat* OR dysfunction*)))
3. #2 OR #1

4. ALL=(“amyloid beta-peptides” OR “tau Protein*” OR “cerebro-spinal fluid*” OR “cerebrospinal fluid*” OR csf OR “spinal fluid*” )
5. TS=(((blood OR plasma) near/3 (biomarker* OR marker* OR “biological marker*”)))
6. TS=(biomarker* OR marker* OR “biological marker*” OR abeta* OR ab42 OR ab40 OR “amyloid beta” OR “beta amyloid” OR “phospho tau*” OR “total tau*” OR ptau181 OR “phosphorylated tau” OR ptau* OR neurofilament )
7. #4 OR #5 OR #6
8. TS=(Diagnos*)
9. TS=(“cost benefit”)
10. TS=(((clinical OR perceived OR clinician* OR pragmatic OR diagnos*) near/5 (impact OR utility OR useful* OR confidence OR decision* OR benefit*)))
11. #9 OR #10
12. #3 AND #7 AND #8 AND #11
13. TS=(animal or animals or pisces or fish or fishes or catfish or catfishes or sheatfish or silurus or arius or heteropneustes or clarias or gariepinus or fathead minnow or fathead minnows or pimephales or promelas or cichlidae or trout or trouts or char or chars or salvelinus or salmo or oncorhynchus or guppy or guppies or millionfish or poecilia or goldfish or goldfishes or carassius or auratus or mullet or mullets or mugil or curema or shark or sharks or cod or cods or gadus or morhua or carp or carps or cyprinus or carpio or killifish or eel or eels or anguilla or zander or sander or lucioperca or stizostedion or turbot or turbots or psetta or flatfish or flatfishes or plaice or pleuronectes or platessa or tilapia or tilapias or oreochromis or sarotherodon or common sole or dover sole or solea or zebrafish or zebrafishes or danio or rerio or seabass or dicentrarchus or labrax or morone or lamprey or lampreys or petromyzon or pumpkinseed or pumpkinseeds or lepomis or gibbosus or herring or clupea or harengus or amphibia or amphibian or amphibians or anura or salientia or frog or frogs or rana or toad or toads or bufo or xenopus or laevis or bombina or epidalea or calamita or salamander or salamanders or newt or newts or triturus or reptilia or reptile or reptiles or bearded dragon or pogona or vitticeps or iguana or iguanas or lizard or lizards or anguis fragilis or turtle or turtles or snakes or snake or aves or bird or birds or quail or quails or coturnix or bobwhite or colinus or virginianus or poultry or poultries or fowl or fowls or chicken or chickens or gallus or zebra finch or taeniopygia or guttata or canary or canaries or serinus or canaria or parakeet or parakeets or grasskeet or parrot or parrots or psittacine or psittacines or shelduck or tadorna or goose or geese or branta or leucopsis or woodlark or lullula or flycatcher or ficedula or hypoleuca or dove or doves or geopelia or cuneata or duck or ducks or greylag or graylag or anser or harrier or circus pygargus or red knot or great knot or calidris or canutus or godwit or limosa or lapponica or meleagris or gallopavo or jackdaw or corvus or monedula or ruff or philomachus or pugnax or lapwing or peewit or plover or vanellus or swan or cygnus or columbianus or bewickii or gull or chroicocephalus or ridibundus or albifrons or great tit or parus or aythya or fuligula or streptopelia or risoria or spoonbill or platalea or leucorodia or blackbird or turdus or merula or blue tit or cyanistes or pigeon or pigeons or columba or pintail or anas or starling or sturnus or owl or athene noctua or pochard or ferina or cockatiel or nymphicus or hollandicus or skylark or alauda or tern or sterna or teal or crecca or oystercatcher or haematopus or ostralegus or shrew or shrews or sorex or araneus or crocidura or russula or european mole or talpa or chiroptera or bat or bats or eptesicus or serotinus or myotis or dasycneme or daubentonii or pipistrelle

or pipistrellus or cat or cats or felis or catus or feline or dog or dogs or canis or canine or canines or otter or otters or lutra or badger or badgers or meles or fitchew or fitch or foumart or foulmart or ferrets or ferret or polecat or polecats or mustela or putorius or weasel or weasels or fox or foxes or vulpes or common seal or phoca or vitulina or grey seal or halichoerus or horse or horses or equus or equine or equidae or donkey or donkeys or mule or mules or pig or pigs or swine or swines or hog or hogs or boar or boars or porcine or piglet or piglets or sus or scrofa or llama or llamas or lama or glama or deer or deers or cervus or elaphus or cow or cows or bos taurus or bos indicus or bovine or bull or bulls or cattle or bison or bisons or sheep or sheeps or ovis aries or ovine or lamb or lambs or mouflon or mouflons or goat or goats or capra or caprine or chamois or rupicapra or leporidae or lagomorpha or lagomorph or rabbit or rabbits or oryctolagus or cuniculus or laprine or hares or lepus or rodentia or rodent or rodents or murinae or mouse or mice or mus or musculus or murine or woodmouse or apodemus or rat or rats or rattus or norvegicus or guinea pig or guinea pigs or cavia or porcellus or hamster or hamsters or mesocricetus or cricetulus or cricetus or gerbil or gerbils or jird or jirds or meriones or unguiculatus or jerboa or jerboas or jaculus or chinchilla or chinchillas or beaver or beavers or castor fiber or castor canadensis or sciuridae or squirrel or squirrels or sciurus or chipmunk or chipmunks or marmot or marmots or marmota or suslik or susliks or spermophilus or cynomys or cottonrat or cottonrats or sigmodon or vole or voles or microtus or myodes or glareolus or primate or primates or prosimian or prosimians or lemur or lemurs or lemuridae or loris or bush baby or bush babies or bushbaby or bushbabies or galago or galagos or anthropoidea or anthropoids or simian or simians or monkey or monkeys or marmoset or marmosets or callithrix or cebuella or tamarin or tamarins or saguinus or leontopithecus or squirrel monkey or squirrel monkeys or saimiri or night monkey or night monkeys or owl monkey or owl monkeys or douroucoulis or aotus or spider monkey or spider monkeys or ateles or baboon or baboons or papio or rhesus monkey or macaque or macaca or mulatta or cynomolgus or fascicularis or green monkey or green monkeys or chlorocebus or vervet or vervets or pygerythrus or hominoidea or ape or apes or hylobatidae or gibbon or gibbons or siamang or siamangs or nomascus or symphalangus or hominidae or orangutan or orangutans or pongo or chimpanzee or chimpanzees or pan troglodytes or bonobo or bonobos or pan paniscus or gorilla or gorillas or troglodytes)

14. #12 NOT #13

**Supplementary Figure 1**

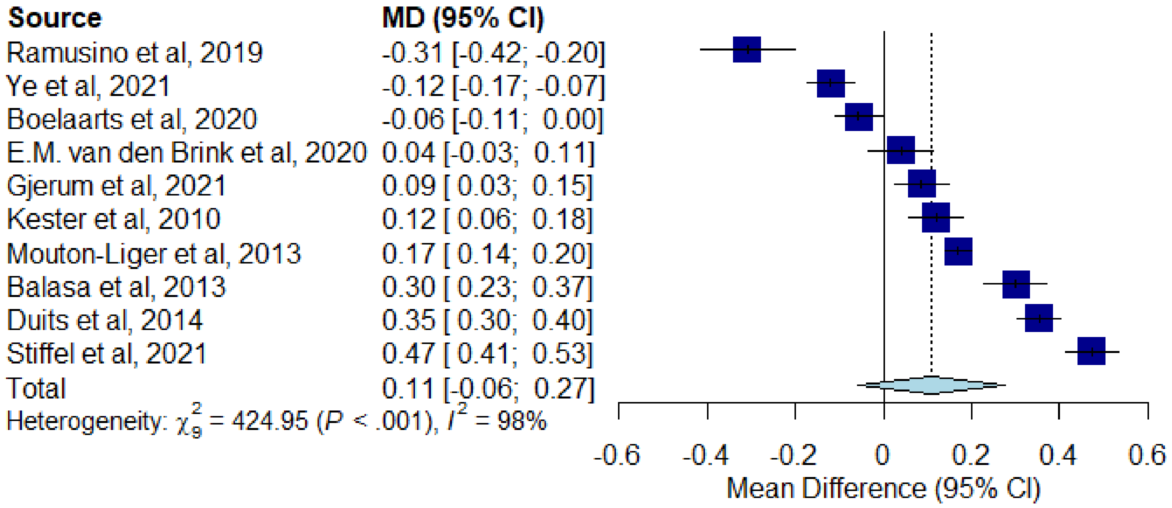


**Fig. 1. (A) Forest plot showing the pooled percentage change from initial AD to final non-AD diagnosis.**

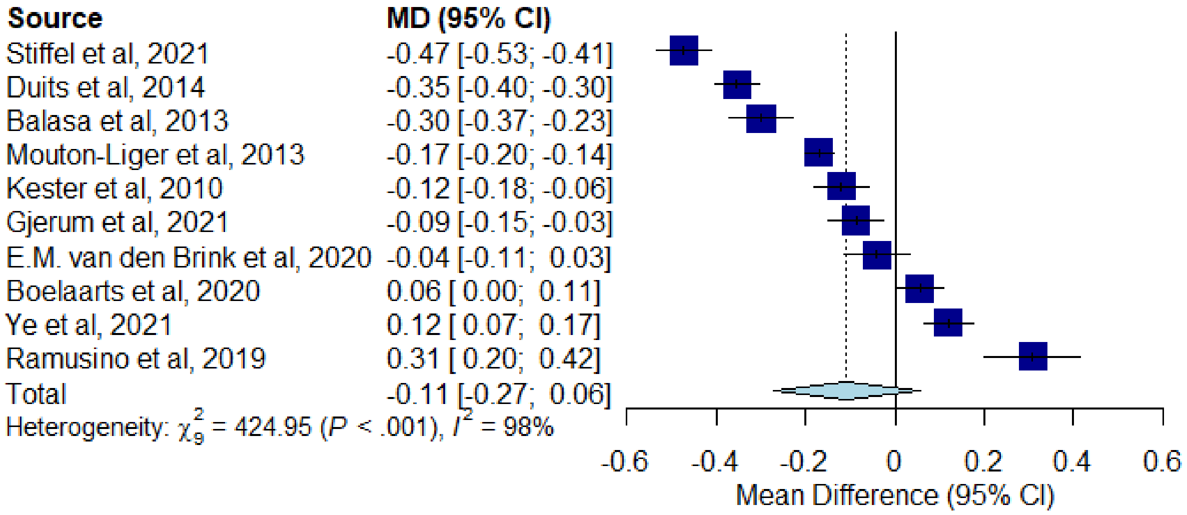


**Fig. 1. (B) Forest plot showing the pooled percentage change from initial non-AD to final AD diagnosis.**

**Supplementary Table 1: Quality Assessment Table**

| Author & Date | Title | Selection bias | Study design | Confounders | Data collection method | Withdrawals & dropouts | Global Rating 1=strong 2=moderate 3=weak |
|---|---|---|---|---|---|---|---|
| **Balasa et al, 2013** | Usefulness of biomarkers in the diagnosis and prognosis of early-onset cognitive impairment | 2 | 2 | 2 | 3 | 1 | 2 |
| **Boelaarts et al, 2020** | Diagnostic Impact of CSF Biomarkers in a Local Hospital Memory Clinic Revisited | 1 | 2 | 2 | 3 | 1 | 2 |
| **Cognat et al, 2019** | What is the clinical impact of cerebrospinal fluid biomarkers on final diagnosis and management in patients with mild cognitive impairment in clinical practice? Results from a nation-wide prospective survey in France | 1 | 2 | 2 | 2 | 1 | 2 |
| **Duits et al, 2014** | Diagnostic impact of CSF biomarkers for Alzheimer's disease in a tertiary memory clinic | 1 | 2 | 2 | 3 | 1 | 2 |
| **E.M. van den Brink et al, 2020** | Clinical impact of CSF assessment on diagnostic accuracy in atypical dementias in Quebec, Canada: preliminary results from a specialized dementia clinic | 1 | 2 | 2 | 3 | 1 | 2 |
| **Falgas et al, 2019** | Clinical applicability of diagnostic biomarkers in early-onset cognitive impairment | 1 | 2 | 2 | 3 | 1 | 2 |

| | | | | | | | |
|---|---|---|---|---|---|---|---|
| **Gjerum et al, 2021** | Comparison of the clinical impact of 2-[18F]FDG-PET and cerebrospinal fluid biomarkers in patients suspected of Alzheimer's disease | 1 | 2 | 2 | 2 | 1 | 2 |
| **Gooblar et al, 2015** | The influence of cerebrospinal fluid (CSF) biomarkers on clinical dementia evaluations | 2 | 2 | 2 | 3 | 1 | 2 |
| **Handels et al, 2017** | Cost-Utility of Using Alzheimer's Disease Biomarkers in Cerebrospinal Fluid to Predict Progression from Mild Cognitive Impairment to Dementia | 1 | 1 | 1 | 2 | 1 | 1 |
| **Keester et al, 2010** | Diagnostic impact of CSF biomarkers in a local hospital memory clinic | 1 | 2 | 2 | 2 | 1 | 2 |
| **Lee et al, 2017** | Cost-effectiveness of cerebrospinal biomarkers for the diagnosis of Alzheimer's disease | 1 | 1 | 1 | 2 | 1 | 1 |
| **Mouton-Liger et al, 2013** | Impact of cerebro-spinal fluid biomarkers of Alzheimer's disease in clinical practice: a multicentric study | 1 | 2 | 2 | 2 | 1 | 2 |
| **Paquet et al, 2016** | Utility of CSF biomarkers in psychiatric disorders: a national multicentre prospective study | 1 | 2 | 2 | 2 | 1 | 2 |
| **Ramusino et al, 2019** | The incremental value of amyloid PET versus CSF biomarkers for the diagnosis of Alzheimer's Disease | 1 | 1 | 2 | 2 | 1 | 2 |

| | | | | | | | |
|---|---|---|---|---|---|---|---|
| **Stiffel et al, 2021** | Use of Alzheimer's Disease Cerebrospinal Fluid Biomarkers in A Tertiary Care Memory Clinic | 2 | 2 | 2 | 2 | 1 | 2 |
| **Valc´arcel-Nazco et al, 2014** | Cost-effectiveness of the use of biomarkers in cerebrospinal fluid for Alzheimer's disease | 1 | 1 | 1 | 1 | 1 | 1 |
| **Willemse et al, 2021** | A neurologist's perspective on serum neurofilament light in the memory clinic: a prospective implementation study | 2 | 2 | 2 | 1 | 1 | 2 |
| **Ye et al, 2021** | Application of Cerebrospinal Fluid AT(N) Framework on the Diagnosis of AD and Related Cognitive Disorders in Chinese Han Population | 2 | 2 | 2 | 2 | 1 | 2 |
